# Do More Appointments Lead to Shorter Waits and Better Patient Experience? A Retrospective Observational Study of NHS Primary Care

**DOI:** 10.64898/2026.07.29.26359221

**Authors:** Rohan Joseph, Harsh Gupta, Shaun Danielli

## Abstract

**Objectives:** To examine whether increased appointment volumes were associated with shorter waiting times or lower patient dissatisfaction, and whether shorter waiting times were associated with lower patient dissatisfaction, in National Health Service (NHS) primary care in England.

**Design:** Retrospective observational study using the Appointments in General Practice dataset and the General Practice Patient Survey (GPPS). Changes were compared across Integrated Care Boards (ICBs) over 2018-2022 and 2022-2023, stratified by staff group and appointment mode, alongside annual panel fixed-effects models.

**Setting:** NHS primary care in England, 2018-2024.

**Participants:** All 42 ICBs in England (294 ICB-year observations).

**Primary Outcome Measures:** Mean waiting time between booking and appointment, and patient dissatisfaction: the share of GPPS respondents rating their overall experience of their GP practice as “neither good nor poor”, “fairly poor” or “very poor”.

**Results:** Completed appointments rose over 2018-2024, with care shifting towards non-GP staff and virtual delivery. Per million additional appointments, waiting time changed by −0.04 days (95% CI −0.10 to 0.03) in 2018-2022 and −0.31 days (−0.57 to −0.04) in 2022-2023, and dissatisfaction by 0.02 percentage points (−0.36 to 0.40) and −0.75 percentage points (−3.05 to 1.56). Per additional day of waiting, dissatisfaction changed by −1.40 percentage points (−3.21 to 0.42) in 2018-2022 and 2.85 percentage points (1.20 to 4.51) in 2022-2023, when changes in appointment volume were small. In fixed-effects models over 2018-2024, all three associations had 95% CIs including zero.

**Conclusions:** Growth in appointment volume did not translate into consistent improvement in waiting times or patient dissatisfaction, and shorter waiting times were not consistently associated with lower dissatisfaction. Either these quality metrics do not respond to output, the key factor under providers’ control, or they do not capture the dimensions of quality that matter. Performance frameworks that assess primary care productivity through these metrics should be reviewed.

**Strengths and Limitations of this Study:**

- The analysis covers all 42 ICBs in England from 2018 to 2024 (294 ICB-year observations), so it includes the full population rather than a sample.
- Cross-ICB comparisons over two periods were complemented by panel fixed-effects models with ICB and year fixed effects, with standard errors clustered at the ICB level.
- Patient dissatisfaction was constructed from harmonised, weighted GPPS response counts, using a question and definition that are consistent across all survey years.
- The analysis is observational and conducted at the ICB level, so associations are not causal and within-area variation across practices and patient groups is not captured.
- Waiting times were derived from administrative categories rather than exact waits, and the analysis includes no direct measures of patient health.

## Introduction

Strengthening primary healthcare is central to improving population health, advancing universal health coverage, and building resilient health systems.^1^ Yet evaluating primary care performance is difficult. Healthcare is not traded on markets, so prices cannot be used to evaluate value. Health benefits are also difficult to incorporate as they accrue over long horizons and are hard to attribute to specific services.^2^ ^3^

In the absence of direct measures of value, key stakeholders, importantly the National Health Service (NHS), rely on alternative, more visible metrics. The first measure is output as captured by appointment volume. In addition, waiting times and patient satisfaction are used for quality adjustments. Each is regularly reported, easy to communicate, and consistently invoked in debates about access to general practice.^4^ ^5^

Public discourse often assumes that quality can be improved by increasing output. In February 2024, for example, NHS England reported that GP practices were delivering two million more appointments per month than before the pandemic. They further described this increase as evidence of progress under its Primary Care Access Recovery Plan.^6^ More appointments should ease constraints on access. Better access should lead to shorter waits. Shorter waits should improve patient satisfaction. Therefore, increased output, whether by providing more inputs or increasing productive efficiency, ought to improve quality.

However, the output-quality relationship is more ambiguous and merits empirical validation. First, waiting times reflect demand as well as supply, so more appointments need not produce shorter waits. If demand rises alongside supply, or if workforce and organisational constraints bind, additional appointments may increase rather than reduce waits.^7^ Second, patient satisfaction is shaped by continuity, communication, expectations, and consultation mode, not just waiting times. Shorter waits need not improve satisfaction, especially if they crowd out other dimensions that patients value. Reported satisfaction also reflects patient characteristics and case mix, so it does not provide a clean signal of provider performance.^8^ ^9^ Many of these factors are beyond the control of the healthcare system, raising questions about the appropriateness of patient satisfaction as a measure of primary care productivity.

We provide a novel empirical analysis of the relationship between appointments, waiting times, and patient satisfaction for NHS primary care in England. Using the Appointments in General Practice and the General Practice Patient Survey, we provide evidence addressing four questions. First, how did appointment volume and the composition of care change over the period? Second, were Integrated Care Boards (ICB) with larger increases in appointment volume those with larger reductions in waiting times? Third, were ICBs with larger increases in appointment volume those with larger reductions in patient dissatisfaction? Fourth, were ICBs with larger reductions in waiting times those with larger reductions in patient dissatisfaction?

## Methods

### Study Design and Setting

We conducted a retrospective observational study of primary care activity, waiting times, and patient dissatisfaction in NHS primary care in England between 2018 and 2024. The unit of analysis was the Integrated Care Board (ICB), the regional administrative body responsible for planning and commissioning NHS services, including primary care.

The analysis examined whether output growth was associated with changes in waiting times and patient satisfaction. We first described national changes in the scale and composition of completed appointments. We then examined the associations between changes in appointment volume and changes in waiting times, between changes in appointment volume and changes in patient dissatisfaction, and between changes in waiting times and changes in patient dissatisfaction across ICBs. These cross-ICB analyses were conducted over two comparison periods: 2018-2022 and 2022-2023. In addition, fixed-effects regression models were estimated.

All analyses were descriptive and correlational. All 42 ICBs in England were included for each year from 2018 to 2024, so the analysis covers the full population of ICBs rather than a sample, yielding 294 ICB-year observations. This study is reported in line with the STROBE guideline.

### Data Sources

#### Appointments in General Practice

Appointment activity data is derived from NHS England’s Appointments in General Practice publications. The data ranges from January 2017 to December 2024. These monthly data report appointments by status, appointment mode, healthcare professional type, and waiting time. We include completed appointments only. Key variables included are total appointment volume, appointment mode, staff type, and waiting time.

Earlier observations were reported at Clinical Commissioning Group (CCG) level rather than ICB level. We mapped CCGs to ICBs using administrative tables (See Appendix C). Monthly data were then aggregated to annual ICB totals for the main analyses.

#### General Practice Patient Survey

Patient dissatisfaction data is obtained from the General Practice Patient Survey (GPPS), covering survey years 2017 to 2025. The analysis panel retains survey years 2019 to 2025, corresponding to analysis years 2018 to 2024 after alignment. Earlier CCG-level results were aggregated to ICB level using the same mapping as described above.

Two GPPS variables were used. The main analysis uses overall experience of the GP practice, which asks respondents to rate their experience on a five-point scale from “very good” to “very poor.” Dissatisfaction is defined as the combined share answering: “neither good nor poor,” “fairly poor,” or “very poor.” The question wording, response scale, and dissatisfaction definition are consistent across all survey years in the analysis panel, with only the variable codes changed (Table D1).

Supplementary analysis uses satisfaction with appointment waiting times. For survey years 2019 to 2023, this item asks about satisfaction on a six-category scale, and dissatisfaction is defined as the combined share answering: “neither satisfied nor dissatisfied,” “fairly dissatisfied,” or “very dissatisfied.” In survey years 2024 and 2025, the GPPS replaced this question with a three-category item on perceived wait adequacy, representing a construct change. This variable is therefore used only as a supplementary harmonised proxy and not as a strictly comparable continuation of the earlier item (Table D2). However, the two measures track each other closely across ICB-year observations (Figure A1).

For both variables, the analysis uses weighted response counts rather than published percentages. Dissatisfaction shares were calculated as weighted dissatisfied responses divided by weighted total responses. Additional detail on GPPS harmonisation, including the crosswalk methodology and weighting, is provided in Appendix D.

#### Alignment of Time Scales

The two datasets were reported on different time bases: appointments by calendar year and GPPS by survey year. GPPS survey years were mapped to analysis years as survey year minus one to reflect the year in which care was provided, so that survey year 2023 corresponds to analysis year 2022. Appointment data were already reported by calendar year and required no adjustment.

### Measures

The main output measure was the annual number of completed appointments within each ICB. We examined total completed appointments, appointments by provider type, and appointments by appointment mode. We also calculated the share of appointments delivered by non-GP staff and the share delivered virtually, defined as telephone and video appointments combined.

Waiting times were measured as the average time between booking and appointment. NHS appointment data report waiting-time categories rather than exact appointment-level waits. Each category was assigned a numeric value in days, as shown in Table E1. Rows in either unknown waiting-time category were not assigned a value and were excluded from both the numerator and the denominator of the waiting-time calculation. These rows are retained in the appointment volume measure used in Figure 1 and in the cross-sectional analyses and are excluded from the panel fixed-effects analyses. They account for less than 0.1% of completed appointments in every year of the sample period.

**Figure 1.**
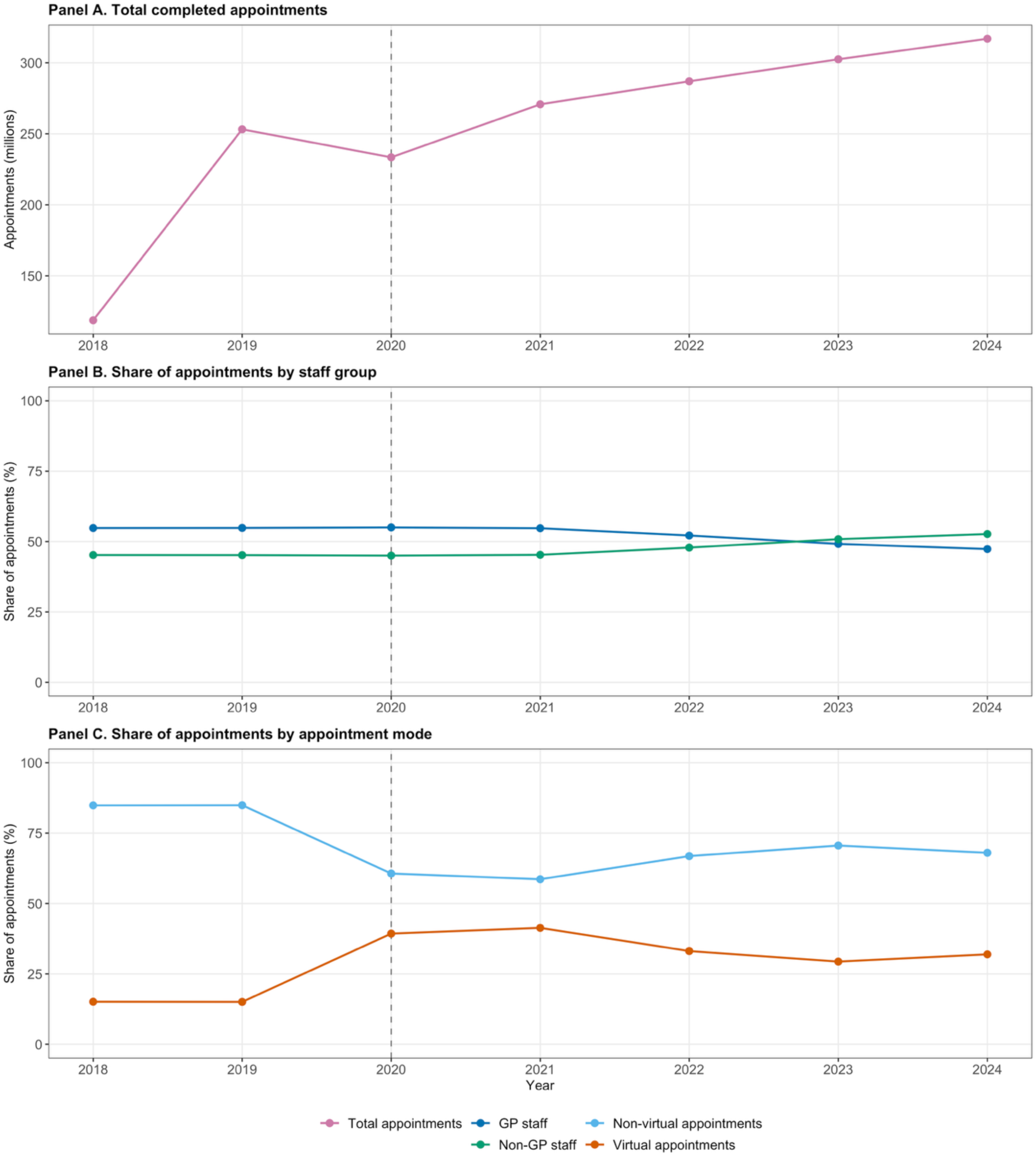
Trends in Primary Care Activity and Care Mix, 2018-2024. *Notes:* Panel A shows annual completed appointment volume, measured in millions. Panels B and C show shares of completed appointments, measured in percent. Panel B separates GP and non-GP sta<; Panel C separates non-virtual and virtual appointments. Virtual appointments combine telephone and video/online appointments; non-virtual appointments are face-to-face appointments. The dashed vertical line marks 2020, the onset of the COVID-19 period.

Mean waiting time was then calculated as the appointment-weighted mean within each ICB-year cell, using completed appointment counts within each waiting-time category as weights. Waiting times were first calculated separately by staff group and appointment mode. Where pooled measures were needed, waiting times were calculated as appointment-weighted averages so that each ICB’s overall waiting-time measure reflected its appointment mix.

Patient dissatisfaction was measured as the share of GPPS respondents rating their overall primary care experience as “very poor”, “fairly poor”, or “neither good nor poor”. Dissatisfaction with waiting times was used in supplementary analysis. The data are aggregated at ICB level and do not permit stratification by patient sex or gender.

### Analytical Approach

The main analysis proceeded in four steps. First, we described national annual trends in completed appointments, the share delivered by non-GP staff, and the share delivered virtually between 2018 and 2024. Second, we examined whether changes in output were associated with changes in waiting times across ICBs. Third, we examined whether changes in appointment volume were directly associated with changes in patient dissatisfaction across ICBs. Fourth, we examined whether changes in waiting times were associated with changes in patient dissatisfaction across ICBs.

These cross-ICB comparisons were made over two periods, 2018-2022 and 2022-2023, stratified by staff group (GP versus non-GP) and by appointment mode (in-person versus virtual).

Ordinary least squares regressions were estimated to summarise and quantify the fitted relationships shown in the figures. We further estimated annual panel fixed-effects models with ICB and year fixed effects. The three specifications are:

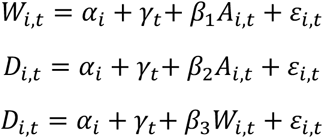

where *A_i_*_,*t*_ is total completed appointments, *W_i_*_,*t*_ is mean waiting time, and *D_i_*_,*t*_ is overall patient dissatisfaction for ICB *i* in year *t*. *α_i_* and *γ_t_* denote ICB and year fixed effects respectively.

For all regression models, standard errors were clustered at the ICB level. Measurement, coverage, and harmonisation choices made to limit bias are described in the Data Sources and Measures sections, and year fixed effects absorb shocks common to all ICBs.

### Patient and Public Involvement

Patients and members of the public were not directly involved in the design or conduct of this study. The analysis relied exclusively on publicly available administrative data and secondary survey data. Patient-reported measures derived from the GPPS were incorporated as key outcome indicators.

## Results

### Changes in Output Volume and Care Mix

In Figure 1, we present aggregate trends in primary care appointment volumes and care mix. Data coverage in the Appointments in General Practice dataset was incomplete in 2018, so absolute volume comparisons are reported from 2019 onwards.

Total completed appointments fell from 253.2 million in 2019 to 233.5 million in 2020 during the COVID-19 pandemic, before rising in each subsequent year to 316.9 million in 2024 (Panel A). Growth was concentrated among non-GP staff. Non-GP appointments rose from 114.4 million in 2019 to 166.9 million in 2024. GP appointments rose to approximately 150 million by 2022 and then remained broadly flat. By 2023, non-GP appointments exceeded GP appointments in absolute terms, and by the end of the study period a majority (52%) of all completed appointments were delivered by non-GP staff (Panel B). Virtual appointments rose sharply during the pandemic, reaching 40% of all completed appointments, before partially reverting to 30% by the end of the study period (Panel C).

### Appointment Volume and Waiting Times

In Figure 2, we present the relationship between changes in output and changes in waiting times across ICBs.

**Figure 2.**
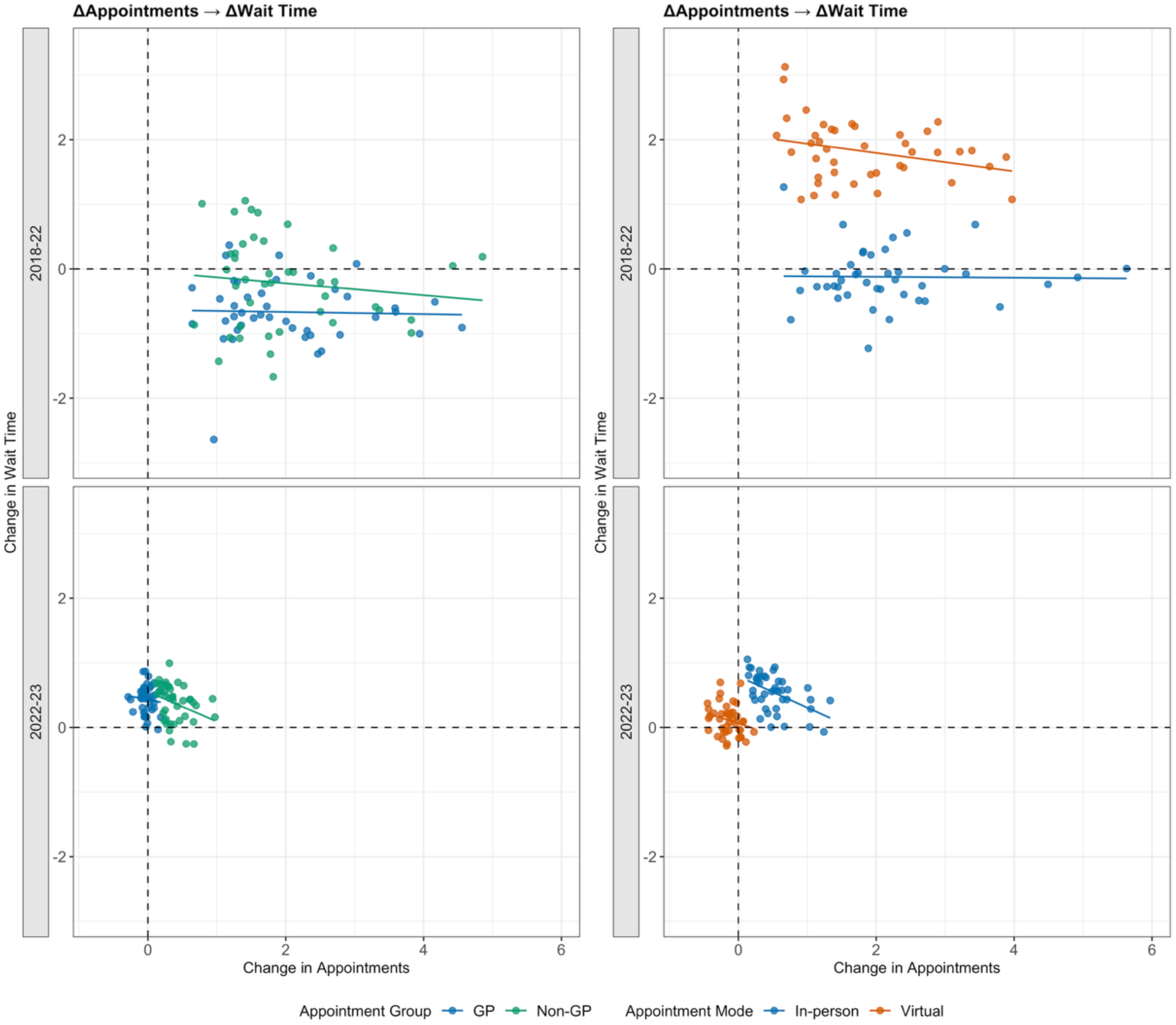
Changes in Appointment Volume and Waiting Time Across ICBs, 2018-2022 and 2022-2023. *Notes:* Panels show ICB-level changes in appointment volume and mean waiting time over 2018-2022 and 2022-2023. Appointment changes are measured in millions of appointments and waiting-time changes are measured in days. The left-hand panels separate GP and non-GP appointments; the right-hand panels separate in-person and virtual appointments. Solid lines show within-period OLS fits. Dashed lines indicate zero change on each axis.

Between 2018-2022, changes in appointment volume were not associated with changes in waiting times. Confidence intervals included zero for all subgroups. The estimated coefficients are: −0.02 days per million additional appointments for GP (95% CI: −0.17, 0.14), −0.09 for non-GP (95% CI: −0.30, 0.12), −0.01 for in-person (95% CI: −0.13, 0.12), and −0.14 for virtual (95% CI: −0.29, 0.01) (Table B1). ICBs with larger increases in appointments did not record larger decreases in waiting times.

Between 2022-2023, changes in output across ICBs were small, so estimates over this period rest on limited variation. Subgroup estimates are: −0.21 for GP (95% CI: −0.82, 0.41), −0.46 for non-GP (95% CI: −0.84, −0.08), −0.48 for in-person (95% CI: −0.74, −0.23), and −0.35 for virtual (95% CI: −0.74, 0.05) (Table B1).

Pooling across staff groups and appointment modes, the association between changes in appointment volume and changes in waiting time was −0.04 days per million additional appointments (95% CI: −0.10, 0.03) in 2018-2022 and −0.31 (95% CI: −0.57, −0.04) in 2022-2023 (Table B1).

### Appointment Volume and Patient Dissatisfaction

In Figure 3, we present the relationship between changes in appointment volume and changes in overall patient dissatisfaction across ICBs.

**Figure 3.**
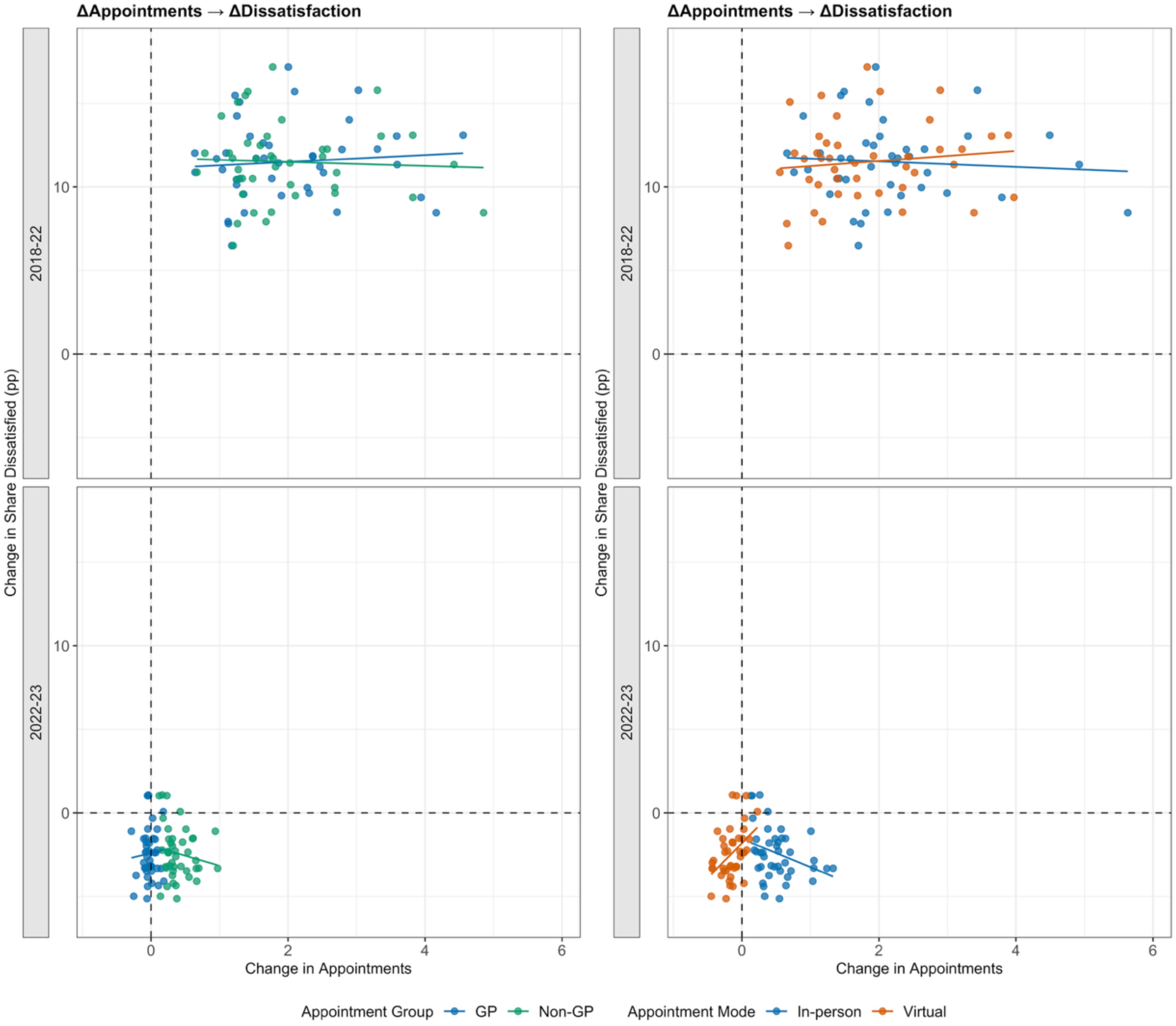
Changes in Appointment Volume and Patient Dissatisfaction across ICBs, 2018-2022 and 2022-2023. *Notes:* Panels show ICB-level changes in appointment volume and overall patient dissatisfaction over 2018-2022 and 2022-2023. Appointment changes are measured in millions of appointments, and dissatisfaction changes are measured in percentage points. The left-hand panels separate GP and non-GP appointments; the right-hand panels separate in-person and virtual appointments. Solid lines show within-period OLS fits. Dashed lines indicate zero change on each axis.

Between 2018-2022, changes in appointment volume were not associated with changes in patient dissatisfaction. Confidence intervals included zero for all subgroups. The estimated coefficients are: 0.20 percentage points per million additional appointments for GP (95% CI: −0.52, 0.92), −0.12 for non-GP (95% CI: −0.86, 0.62), −0.16 for in-person (95% CI: −0.79, 0.46), and 0.30 for virtual (95% CI: −0.48, 1.08) (Table B2).

Between 2022-2023, subgroup estimates diverged by appointment mode. More in-person appointments were associated with lower dissatisfaction: −1.69 percentage points per million additional appointments (95% CI: −3.08, −0.30) (Table B2). More virtual appointments were associated with higher dissatisfaction: 4.17 (95% CI: 1.94, 6.40) (Table B2). Staff-group estimates showed no association: 0.99 for GP (95% CI: −4.44, 6.41) and −1.21 for non-GP (95% CI: −3.69, 1.27) (Table B2).

Pooling across staff groups and appointment modes, the direct association between changes in appointment volume and changes in dissatisfaction was 0.02 percentage points per million additional appointments (95% CI: −0.36, 0.40) in 2018-2022 and −0.75 (95% CI: −3.05, 1.56) in 2022-2023 (Table B2).

### Waiting Times and Patient Dissatisfaction

In Figure 4, we present the relationship between changes in waiting times and changes in overall patient dissatisfaction across ICBs.

**Figure 4.**
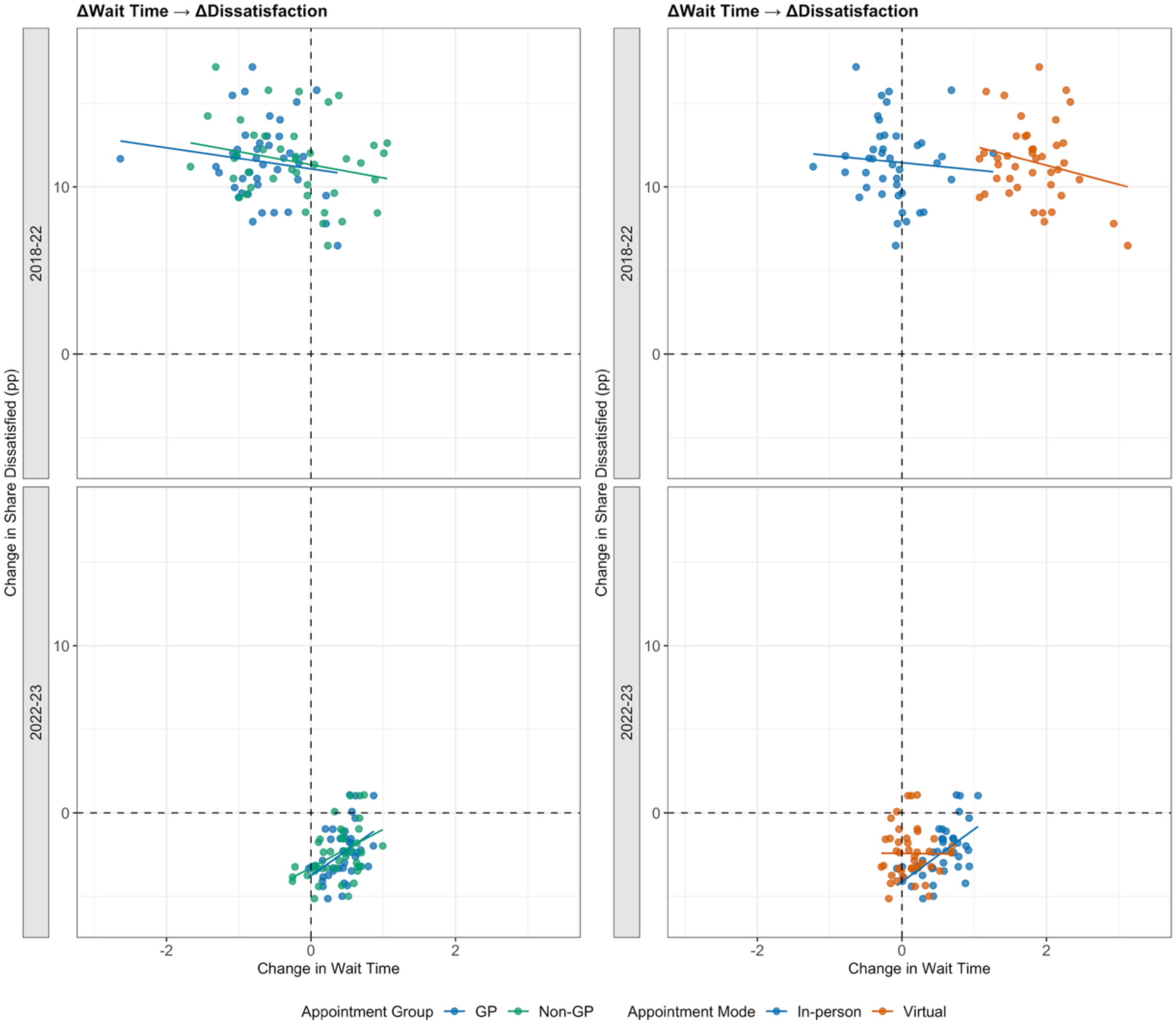
Changes in Waiting Time and Patient Dissatisfaction across ICBs, 2018-2022 and 2022-2023. *Notes:* Panels show ICB-level changes in mean waiting time and overall patient dissatisfaction over 2018-2022 and 2022-2023. Waiting-time changes are measured in days, and dissatisfaction changes are measured in percentage points. The left-hand panels separate GP and non-GP waiting times; the right-hand panels separate in-person and virtual waiting times. Solid lines show within-period OLS fits. Dashed lines indicate zero change on each axis.

Between 2018-2022, changes in waiting times were not associated with changes in patient dissatisfaction. Confidence intervals included zero for all subgroups. The estimated coefficients are: −0.63 percentage points per additional day for GP (95% CI: −2.12, 0.85), −0.79 for non-GP (95% CI: −1.79, 0.22), −0.43 for in-person (95% CI: −1.88, 1.03), and −1.14 for virtual (95% CI: −2.94, 0.66) (Table B3).

Between 2022-2023, a positive relationship emerged for non-virtual subgroups. The estimated coefficients are: +3.06 percentage points per additional day for GP (95% CI: 1.15, 4.97), +2.30 for non-GP (95% CI: 1.01, 3.58), and +3.15 for in-person (95% CI: 1.74, 4.56) (Table B3). The virtual subgroup showed no relationship: −0.05 (95% CI: −1.65, 1.56) (Table B3).

Pooling across staff groups and appointment modes, the association between changes in waiting time and changes in dissatisfaction was −1.40 percentage points per additional day (95% CI: −3.21, 0.42) in 2018-2022 and 2.85 (95% CI: 1.20, 4.51) in 2022-2023 (Table B3).

### Panel Fixed-Effects Estimates

Table 1 reports annual panel fixed-effects estimates using within-ICB variation over the full 2018-2024 period. Appointment volume was weakly associated with waiting time (−0.074 days per million appointments, 95% CI: −0.156, 0.007), with the interval including zero. Appointment volume was not associated with overall dissatisfaction (−0.105 percentage points per million appointments, 95% CI: −0.389, 0.178). Waiting time was not associated with overall dissatisfaction (0.319 percentage points per additional day, 95% CI: −0.387, 1.025).

**Table 1.** Annual Panel Fixed-Effects Estimates of Appointment Volume, Waiting Time, and Overall Dissatisfaction, 2018-2024.

|  | **(1) Wait time** | **(2) Overall dissatisfaction** | **(3) Overall dissatisfaction** |
| --- | --- | --- | --- |
| Appointment volume<br>(millions) | -0.0743*<br>(0.0404) | -0.1055<br>(0.1402) |  |
| Waiting time<br>(days) |  |  | 0.3190<br>(0.3496) |
| ICB fixed effects | Yes | Yes | Yes |
| Year fixed effects | Yes | Yes | Yes |
| Observations | 294 | 294 | 294 |
| R<sup>2</sup> (Overall) | 0.9473 | 0.9624 | 0.9625 |

\*\*\* p < 0.01, \*\* p < 0.05, \* p < 0.1
Notes: Annual ICB-level fixed-effects estimates for 2018-2024. Column 1 regresses waiting time on appointment volume; column 2 regresses overall dissatisfaction on appointment volume; column 3 regresses overall dissatisfaction on waiting time. Appointment volume is measured in millions, waiting time in days, and overall dissatisfaction in percentage points. All models include ICB and year fixed effects. SEs clustered at ICB level are shown in parentheses. ICB, Integrated Care Board.

## Discussion

Between 2018 and 2024, primary care output in England increased and the composition of care shifted towards non-GP staff and virtual delivery. Across ICBs, changes in appointment volume were not meaningfully associated with changes in waiting times. Estimated relationships were indistinguishable from zero in 2018-2022 and small in 2022-2023. Appointment volume was also not directly associated with patient dissatisfaction. Changes in waiting times were not associated with changes in patient dissatisfaction in 2018-2022. A positive association emerged in 2022-2023 for non-virtual subgroups but was absent for virtual appointments. Annual panel fixed-effects models exploiting within-ICB variation over the full 2018-2024 period yield the same conclusion, with all three intervals including zero. Output growth did not translate into consistent improvement on quality metrics.

Waiting times reflect demand as well as supply. If demand rises alongside supply, additional appointments absorb existing pressure rather than reduce waits.^7^ The composition of care also changed over the period. Appointment growth coincided with a rising share of non-GP and virtual delivery. A higher appointment count did not represent the same type of care over time. Waiting times may also respond to dissatisfaction rather than drive it, if practices facing poorer reported experience alter how they schedule appointments.

Patient satisfaction is similarly shaped by more than waiting times. Continuity of care, communication, consultation mode, and patient expectations all contribute to reported experience.^8^ ^9^ Satisfaction also reflects patient characteristics and case mix, which do not indicate provider performance.^9^ This is consistent with dissatisfaction responding to dimensions of care that waiting times do not capture. The direct relationship between appointment volume and dissatisfaction reinforces this. In 2022-2023, more in-person appointments were associated with lower dissatisfaction while more virtual appointments were associated with higher dissatisfaction. Dissatisfaction responded differently depending on how care was delivered, not just how much was delivered.

These findings have two possible interpretations, both with the same practical implication. First, waiting times and patient satisfaction may be genuinely informative but driven by factors other than appointment volume. Demand conditions, workforce composition, continuity, and case mix may all play a role. If so, the priority is to understand what actually moves these metrics. Second, these metrics may not adequately capture the quality dimensions that matter for primary care. Neither the output measure nor the quality adjustments used in current frameworks incorporate direct measures of patient health. Such measures are difficult to attribute to individual GP appointments but are ultimately the outcome that primary care is intended to improve. These results are consistent with wider work showing that activity-based measures can diverge from indicators of quality or benefit.^10–13^ Under either interpretation, evaluating primary care productivity through output adjusted by waiting times and patient satisfaction should be reconsidered.

This study has several limitations. The analysis is observational and descriptive. The reported associations should not be interpreted causally. The direction of association between waiting times and dissatisfaction cannot be established from these data. It is conducted at ICB level, so it cannot capture within-area variation across practices, patient groups, or clinical need. Waiting times were derived from administrative categories rather than exact appointment-level waits. Category midpoints were assigned as shown in Table E1, and the open-ended category was assigned 30 days. GPPS variables required harmonisation across survey years, as detailed in Appendix D. Data coverage in the appointments dataset was lower in 2018 than in subsequent years, which may affect the magnitude of measured 2018-2022 changes across ICBs. The analysis does not include direct measures of patient health. Attributing health outcomes to individual GP appointments is inherently difficult, but this absence means the paper evaluates quality only through the same metrics whose adequacy it questions.

## Conclusion

In NHS primary care in England between 2018 and 2024, growth in appointment volume did not translate into consistent improvement in waiting times or patient dissatisfaction, and shorter waiting times were not consistently associated with lower dissatisfaction. Either current quality metrics are driven by factors other than appointment volume, or they do not capture the dimensions of patient welfare that matter for primary care. In either case, performance frameworks that evaluate primary care productivity through output in conjunction with waiting times and patient satisfaction should be reviewed. Future research should identify what drives these metrics and assess whether they reflect genuine improvements in patient health.

## Statements

### Contributors

RJ, HG and SD conceived the study. RJ and HG led the data analysis and first draft of the manuscript. All authors contributed to subsequent drafting of the manuscript and accept full responsibility for, and have read and approved, the final manuscript. RJ is the guarantor. The corresponding author attests that all listed authors meet authorship criteria and that no others meeting the criteria have been omitted. The views expressed in this manuscript are those of the authors and not necessarily those of their institutions.

### Patient and Public Involvement

Patients and/or the public were not involved in the design, or conduct, or reporting, or dissemination plans of this research. Refer to the Methods section for further details.

### Patient Consent for Publication

Not applicable.

### Ethics Approval

This study used publicly available, aggregated administrative and survey data with no identifiable patient information. Ethical approval was not required under UK research governance guidelines. No individual patient data were accessed, so the Declaration of Helsinki does not apply.

### Provenance and Peer Review

Not commissioned; externally peer reviewed.

### AI Use

During the preparation of this work, the authors used generative AI (ChatGPT) to check for grammar, punctuation errors, and language refinement. After using this tool, the authors reviewed and edited the text as needed and take full responsibility for the content of the publication.

## Data Availability

All source data used in this study are publicly available. Appointments in General Practice data are published by NHS England and available at https://digital.nhs.uk/data-and-information/publications/statistical/appointments-in-general-practice. General Practice Patient Survey data are published by NHS England and available at https://gp-patient.co.uk.

The CCG-to-ICB crosswalk and the GPPS variable crosswalk were constructed by the authors from public sources as described in Appendices C and D. Both files and the analysis code are available from the corresponding author on request.

## Appendix

## Appendix A. Correlation between Waiting-Time Dissatisfaction and Overall Dissatisfaction

**Figure A1.**
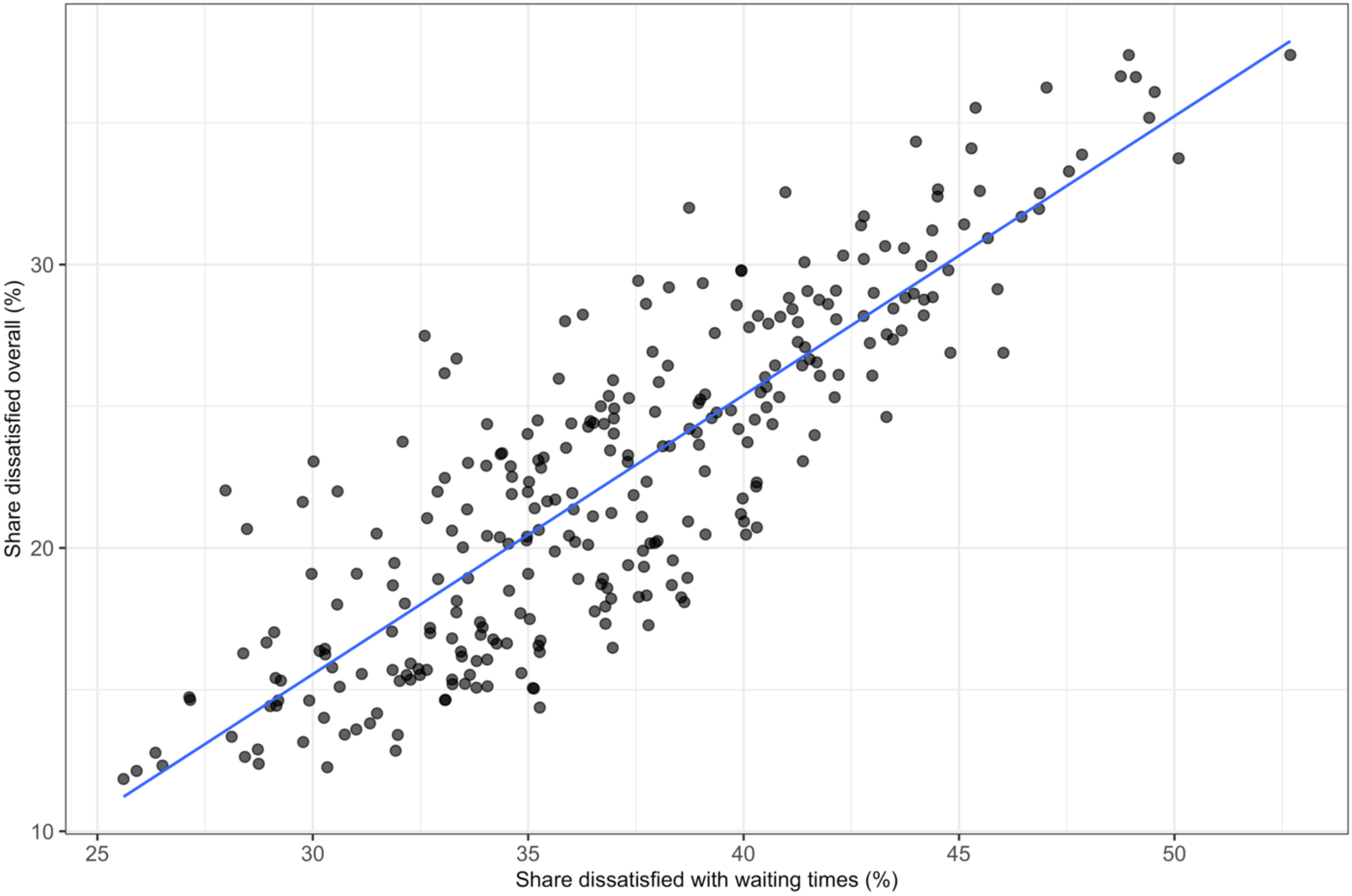
Correlation between Dissatisfaction with Waiting Time and Overall Dissatisfaction across ICB-Year Observations

*Notes:* Each point represents an ICB-year observation from the GP Patient Survey. The horizontal axis shows the percent of respondents classified as dissatisfied with waiting times, and the vertical axis shows the percent classified as dissatisfied with their overall primary care experience. The solid line shows the fitted linear relationship.

## Appendix B. Cross-Sectional Regression Estimates for the Relationships between Appointment Volume, Waiting Time, and Patient Dissatisfaction

**Table B1.** Cross-Sectional Estimates of Changes in Waiting Time on Changes in Appointment Volume.

|  | Staff group |  | Appointment mode |  | Pooled |
| --- | --- | --- | --- | --- | --- |
|  | GP | Non-GP | In-person | Virtual | Total |
| *Dependent variable: Change in waiting time (days)* |  |  |  |  |  |
| **Panel A. 2018-2022** |  |  |  |  |  |
| Change in appointment volume (millions) | -0.0162<br>(0.0760) | -0.0922<br>(0.1049) | -0.0071<br>(0.0631) | -0.1424*<br>(0.0745) | -0.0350<br>(0.0322) |
| Observations | 42 | 42 | 42 | 42 | 42 |
| R<sup>2</sup> | 0.0010 | 0.0159 | 0.0003 | 0.0839 | 0.0249 |
| **Panel B. 2022-2023** |  |  |  |  |  |
| Change in appointment volume (millions) | -0.2087<br>(0.3043) | -0.4609**<br>(0.1873) | -0.4833***<br>(0.1250) | -0.3485*<br>(0.1950) | -0.3082**<br>(0.1311) |
| Observations | 42 | 42 | 42 | 42 | 42 |
| R<sup>2</sup> | 0.0090 | 0.1027 | 0.2579 | 0.0569 | 0.0922 |

\*\*\* p < 0.01, \*\* p < 0.05, \* p < 0.1
*Notes:* Cross-sectional OLS estimates across ICBs. The dependent variable is the change in mean waiting time, measured in days. The independent variable is the change in appointment volume, measured in millions. Panel A reports 2018-2022 changes; Panel B reports 2022-2023 changes. Columns 1-2 show staff group, columns 3-4 appointment mode, and column 5 the pooled estimate. SEs clustered at ICB level are shown in parentheses. ICB, Integrated Care Board.

**Table B2.** Cross-Sectional Estimates of Changes in Patient Dissatisfaction on Changes in Appointment Volume.

|  | Staff group |  | Appointment mode |  | Pooled |
| --- | --- | --- | --- | --- | --- |
|  | GP | Non-GP | In-person | Virtual | Total |
| *Dependent variable: Change in patient dissatisfaction (percentage points)* |  |  |  |  |  |
| **Panel A. 2018-2022** |  |  |  |  |  |
| Change in appointment volume (millions) | 0.2030<br>(0.3557) | -0.1201<br>(0.3652) | -0.1643<br>(0.3102) | 0.3016<br>(0.3875) | 0.0232<br>(0.1887) |
| Observations | 42 | 42 | 42 | 42 | 42 |
| R<sup>2</sup> | 0.0072 | 0.0024 | 0.0055 | 0.0146 | 0.0003 |
| **Panel B. 2022-2023** |  |  |  |  |  |
| Change in appointment volume (millions) | 0.9876<br>(2.6865) | -1.2070<br>(1.2280) | -1.6906**<br>(0.6861) | 4.1713***<br>(1.1050) | -0.7465<br>(1.1405) |
| Observations | 42 | 42 | 42 | 42 | 42 |
| R<sup>2</sup> | 0.0040 | 0.0256 | 0.1085 | 0.1963 | 0.0128 |

\*\*\* p < 0.01, \*\* p < 0.05, \* p < 0.1
Notes: Cross-sectional OLS estimates across ICBs. The dependent variable is the change in overall dissatisfaction, measured in percentage points. The independent variable is the change in appointment volume, measured in millions. Panel A reports 2018-2022 changes; Panel B reports 2022-2023 changes. Columns 1-2 show staff group, columns 3-4 appointment mode, and column 5 the pooled estimate. SEs clustered at ICB level are shown in parentheses. ICB, Integrated Care Board.

**Table B3.** Cross-Sectional Estimates of Changes in Patient Dissatisfaction on Changes in Waiting Time.

|  | Staff group |  | Appointment mode |  | Pooled |
| --- | --- | --- | --- | --- | --- |
|  | GP | Non-GP | In-person | Virtual | Total |
| *Dependent variable: Change in patient dissatisfaction (percentage points)* |  |  |  |  |  |
| **Panel A. 2018-2022** |  |  |  |  |  |
| Change in waiting time (days) | -0.6320<br>(0.7360) | -0.7862<br>(0.4962) | -0.4278<br>(0.7202) | -1.1434<br>(0.8910) | -1.3961<br>(0.8993) |
| Observations | 42 | 42 | 42 | 42 | 42 |
| R<sup>2</sup> | 0.0189 | 0.0559 | 0.0066 | 0.0509 | 0.0625 |
| **Panel B. 2022-2023** |  |  |  |  |  |
| Change in waiting time (days) | 3.0625***<br>(0.9459) | 2.2948***<br>(0.6359) | 3.1486***<br>(0.6972) | -0.0456<br>(0.7948) | 2.8540***<br>(0.8200) |
| Observations | 42 | 42 | 42 | 42 | 42 |
| R<sup>2</sup> | 0.1863 | 0.1912 | 0.3409 | 0.0001 | 0.1925 |

\*\*\* p < 0.01, \*\* p < 0.05, \* p < 0.1
*Notes:* Cross-sectional OLS estimates across ICBs. The dependent variable is the change in overall dissatisfaction, measured in percentage points. The independent variable is the change in mean waiting time, measured in days. Panel A reports 2018-2022 changes; Panel B reports 2022-2023 changes. Columns 1-2 show staff group, columns 3-4 appointment mode, and column 5 the pooled estimate. SEs clustered at ICB level are shown in parentheses. ICB, Integrated Care Board.

## Appendix C. CCG-to-ICB Geographic Mapping

Following NHS England’s transition from Clinical Commissioning Groups (CCGs) to Integrated Care Boards (ICBs), a geographic linkage between legacy CCG identifiers and current ICB geography was required. We constructed this linkage using postcode-level Office for National Statistics Postcode Directory data, combining the August 2018 ONSPD file containing CCG codes with the May 2025 ONSPD file containing current Sub-ICB codes.

The two postcode files were deduplicated and merged on postcode. The resulting postcode overlaps were linked to ONS CCG and Sub-ICB code/name reference files, then aggregated to CCG-by-Sub-ICB cells. For each CCG-to-Sub-ICB pair, we counted matched postcodes and calculated an allocation weight equal to the pair’s postcode count divided by the CCG’s matched postcode count. CCGs spanning multiple current Sub-ICBs were therefore retained as multiple weighted rows rather than being manually assigned to a single destination.

ICB and region identifiers were added from the NHS England appointments Sub-ICB data dictionary, which provides the current Sub-ICB-to-ICB hierarchy. The final geography outputs include a CCG-to-Sub-ICB weighted crosswalk, a Sub-ICB-to-ICB lookup, and a mixed lookup that supports both legacy CCG codes and current Sub-ICB codes in downstream appointment and GP Patient Survey processing.

## Appendix D. GP Patient Survey Harmonisation across Survey Years

The GP Patient Survey (GPPS) was used to construct patient-reported dissatisfaction measures from harmonised weighted response counts. The GPPS harmonisation covers survey years 2017 to 2025, but the analysis panel retains survey years 2019 to 2025. Survey years are aligned to appointment years as: analysis year = survey year −1. Survey years 2019-2025 therefore correspond to analysis years 2018-2024, the appointment-year range used in the main paper. Survey years 2017 and 2018 are harmonised in the pipeline but excluded from the analysis panel because they map to appointment years 2016 and 2017.

Survey results were published at Clinical Commissioning Group (CCG) level for retained survey years 2019-2021 and at Integrated Care Board (ICB) level from survey year 2022 onwards. CCG-level rows were mapped to current ICB geography using the mixed geography lookup described in Appendix C, which allocates legacy CCGs to current Sub-ICB and ICB geographies using postcode-overlap weights before summing weighted response counts to ICB-year-response rows. Survey years 2022-2025 use the published ICB-level geography fields directly.

Variables were aligned across survey years using a manually reviewed master crosswalk anchored on the 2019 GPPS variable dictionary. Survey year 2019 was selected as the base because it is the first GPPS year retained in the analysis panel after aligning survey years to appointment years, and because the preliminary fuzzy name-matching audit across GPPS dictionaries showed that 2019 provided the highest cross-year variable coverage. The crosswalk maps each year’s source variable codes onto the 2019 base variable and response definitions. Blank crosswalk cells are not carried into the harmonised output, and rows identified as percentages, totals, or summaries are excluded from the final count-based output.

The analysis uses weighted response counts rather than published percentage variables. For each ICB-year-question, weighted response counts are summed across the relevant response categories. Dissatisfaction shares are then calculated as weighted dissatisfied responses divided by weighted total responses. No additional survey reweighting or imputation is applied. The only weighting introduced by the pipeline is the geographic allocation weight used to map legacy CCG-level rows to current ICB geography.

Two GPPS variables were used in the analysis. Their harmonisation is described below.

### Overall Experience of GP (Main Analysis)

This variable asks respondents to rate their overall experience of their GP practice. The question wording, response scale, and dissatisfaction definition are consistent across all survey years used in the analysis. Variable codes changed from Q28_1 – Q28_5 (survey years 2019-2023) to overallexp_1.count – overallexp_5.count (survey years 2024-2025), but the underlying item and response categories are unchanged.

Dissatisfaction is defined as the combined share of respondents answering, “neither good nor poor,” “fairly poor,” or “very poor.”

**Table D1.** Response Category Crosswalk for Overall Experience of GP.

| Response Category | 2019-2023 Variable | 2024-2025 Variable | Dissatisfied |
| --- | --- | --- | --- |
| Very good | Q28_1 | overallexp_1.count | No |
| Fairly good | Q28_2 | overallexp_2.count | No |
| Neither good nor poor | Q28_3 | overallexp_3.count | Yes |
| Fairly poor | Q28_4 | overallexp_4.count | Yes |
| Very poor | Q28_5 | overallexp_5.count | Yes |

Notes: Response-count variables used to construct overall patient dissatisfaction. Categories marked dissatisfied form the numerator; all retained response categories form the denominator. Survey years 2019-2025 are mapped to appointment years 2018-2024. Earlier survey years are harmonised in the pipeline but excluded from the retained analysis panel.

### Satisfaction with GP Waiting Times (Supplementary Analysis)

For survey years 2019-2023, this variable asks respondents about their satisfaction with general practice appointment times on a six-category scale. Dissatisfaction is defined as the combined share answering, “neither satisfied nor dissatisfied,” “fairly dissatisfied,” or “very dissatisfied.” The “I’m not sure when I can get an appointment” category is retained in the denominator but not counted as dissatisfied.

In survey years 2024 and 2025, the GPPS replaced this question with a three-category item asking respondents to evaluate how long they waited for their appointment. This is a construct change, from satisfaction with appointment times to perceived adequacy of wait length. The current crosswalk maps this newer item onto the 2019-based Q25 structure, as shown in Table D2. Because the analysis uses the 2019 base response labels after mapping, the 2024-2025 waiting-time dissatisfaction share is calculated as:

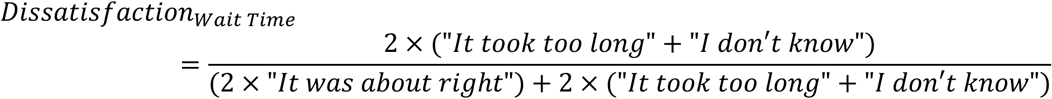

This variable is therefore used only as a supplementary harmonised proxy, not as a strictly comparable continuation of the 2019-2023 satisfaction item.

**Table D2.** Response Category Crosswalk for Satisfaction with Appointment Times.

| **Response Category** | **2019-2023 Variable** | **2024-2025 Variable** | **Dissatisfied** |
| --- | --- | --- | --- |
| Very satisfied | Q25_1 | lastgpapptwait_1.count | No |
| Fairly satisfied | Q25_2 | lastgpapptwait_1.count | No |
| Neither satisfied nor dissatisfied | Q25_3 | lastgpapptwait_3.count | Yes |
| Fairly dissatisfied | Q25_4 | lastgpapptwait_2.count | Yes |
| Very dissatisfied | Q25_5 | lastgpapptwait_2.count | Yes |
| I'm not sure when I can get an appointment | Q25_6 | - | No |

*Notes:* Response-count variables used to construct dissatisfaction with appointment times. Categories marked dissatisfied form the numerator; all retained response categories form the denominator. Survey years 2019-2025 are mapped to appointment years 2018-2024. Q25\_6 is retained in the denominator for 2019-2023 and has no mapped 2024-2025 equivalent.

## Appendix E. Appointments in General Practice Waiting-Time Categories

The value assigned to the open-ended category is discretionary. We chose the smallest value consistent with the category’s lower bound, which minimizes assumptions about the unobserved upper tail of the waiting-time distribution. We then re-estimated the pooled specifications reported in Table 1 and in Tables B1 and B3 using values of 35, 42, and 56 days. Because waiting time enters as the dependent variable in some specifications and as the independent variable in others, raising the assigned value increases the magnitude of the former and reduces the magnitude of the latter. No estimate changed sign and no confidence interval changed its relationship to zero. Full results are available from the corresponding author on request.

**Table E1.** Waiting-Time Category Values Used to Construct Mean Waiting Time.

| Waiting-time Category | Assigned Value in Days |
| --- | --- |
| Same Day | 0 |
| 1 Day | 1 |
| 2 to 7 Days | 4.5 |
| 8 to 14 Days | 11 |
| 15 to 21 Days | 18 |
| 22 to 28 Days | 25 |
| More than 28 Days | 30 |
| Unknown / Data Issue | Excluded |
| Unknown / Data Quality | Excluded |

*Notes:* Values are assigned to Appointments in General Practice waiting-time categories before aggregation to ICB-year cells. Values for bounded categories are category midpoints. "Same Day" is assigned zero days. "More than 28 Days" is open-ended and is assigned 30 days. Rows in either unknown category are assigned no value and are excluded from both the numerator and the denominator of the appointment-weighted mean. These rows are retained in the appointment volume measure used in Figure 1 and in the cross-sectional analyses. ICB, Integrated Care Board.

